# Robustness and bias of European excess death estimates in 2020 under varying model specifications

**DOI:** 10.1101/2021.06.04.21258353

**Authors:** Jonas Schöley

**Author notes:** **Corresponding authors**: Jonas Schöley.

## Abstract

Various procedures are in use to calculate excess deaths during the ongoing COVID-19 pandemic. Using weekly death counts from 20 European countries, we evaluate the robustness of excess death estimates to the choice of model for expected deaths and perform a cross-validation analysis to assess the error and bias in each model’s predicted death counts. We find that the different models produce very similar patterns of weekly excess deaths but disagree substantially on the level of excess. While the exact country ranking along percent excess death in 2020 is sensitive to the choice of model the top and bottom ranks are robustly identified. On the country level, the 5-year average death rate model tends to produce the lowest excess death estimates, whereas high excess deaths are produced by the popular 5-year average death count and Euromomo-style Serfling models. Cross-validation revealed these estimates to be biased under a causal interpretation of “expected deaths had COVID-19 not happened.”

## Introduction

The concept of “excess deaths” became widely known in 2020. Early in the year, international newspapers reported weekly death counts that were notably elevated compared to previous years [30, 33]. In a situation where tests for COVID-19 were scarce and the underlying cause of death often hard to determine, excess deaths proved to be a rational means of monitoring the direct and indirect mortality impact of the ongoing COVID-19 pandemic with minimal data requirements [16]. Since the start of the pandemic, the literature on excess deaths has been growing fast and already features international comparisons and rankings owing to the readily available data on death counts for many countries [14, 12, 22, 4]. However, despite the prominence and relevance of the measure, the excess death methodology is far from standardized with different definitions of excess deaths and different models used to estimate an expected death counts against which the excess is judged. This eclectic state of the art begs the questions on robustness and bias of excess death estimates and cross-country comparisons under various analysis strategies.

Given data from 20 European countries, we will assess how excess death measures in the year 2020 vary under different modeling choices for the expected number of deaths. Furthermore, we test the models for bias in their predicted weekly and annual death counts using a time-series cross-validation setup. While the lack of consensus extends beyond the model specification to the very definition of excess deaths, we will focus entirely on the impact of model specification and define excess as observed minus expected, allowing the measure to be negative.

As the literature on excess deaths in 2020 is dominated by simple average or regression-based approaches, we will restrict our attention to this class of models, focusing on the effect of different specifications. Models based upon multi-year averages are prominently employed by newspapers and statistical offices and featured in the academic literature [5, 17, 18, 20]. While simple and easy to communicate, these procedures do not adjust for time trends in the death counts or rates. This issue is addressed by the Serfling model [27], which in various specifications has been applied to quantify covid related deaths [3, 31, 32, 9, 12]. Generalized additive models relax the strict specifications of the Serfling model and allow for smooth long-term and seasonal effects [1, 26]. These models may be further elaborated via the inclusion of temperature effects, autoregressive residuals, adjustments for bank holidays, and Bayesian inference [14].

Having chosen 9 model specifications reflecting the heterogeneity in the literature (Table 1), we probe each model for a tendency to produce high or low estimate of expected and conversely excess deaths, both weekly and total. We ask if the ranking of European countries along the total percentage of excess deaths in 2020 differs under different baseline models, and we identify those countries where there is disagreement about the existence of excess deaths among the models.

**Table 1:** Models for weekly expected deaths.

| Model | Description | References |
| --- | --- | --- |
| **Weekly averages**<br>(AVGc5) 5 year average death counts<br>(AVGr5) 5 year average death rates | Average over the weekly deaths counts or rates in the preceding 5 years implemented as quasi-Poisson GLM with week-of-year coefficients | [20], [29], [5] |
| **Serfling Model**<br>(SRFc) without exposures<br>(SRFcem) Euromomo style, i.e. no exposures, fitted only on weeks without flu-activity<br>(SRFr) with exposures | Quasi-Poisson regression on death counts with log-linear long term trend, AIC selected Fourier-term seasonality, and public-holiday coefficients | [3], [31], [32], [9], [12] |
| **Generalized Additive Model**<br>(GAMr) without temperature anomaly predictor<br>(GAMrt) with temperature anomaly predictor as smoothly varying coefficient over week of year | Quasi-Poisson regression on death counts with log-linear long term trend, penalized cyclical spline seasonality, and public-holiday coefficients | [1], [26] |
| **Latent Gaussian Model**<br>(LGMr) without temperature anomaly predictor<br>(LGMrt) with temperature anomaly predictor as varying coefficient over week of year implemented as cyclic random walk | Bayesian Poisson regression on death counts with autoregressive trend, non-parametric time-varying seasonality, and public-holiday coefficients | [14] |

When excess death numbers vary by model, the question arises which model to trust. In line with [14] and [19] we argue that the challenge of estimating the expected number of weekly deaths in the year 2020, given that COVID had not occurred, is a classical forecasting challenge, and thus, any model can be validated by testing its predictions on past data. To that end, we construct a cross-validation challenge, where each model is fitted on seven years of weekly death counts and is then tasked to predict the next 52 weeks from that series. The error and bias of any model can then be estimated by comparing the model predictions with the observed data. If a model’s predictions over past years tend to over/underestimate observed death counts, we argue that this model also over/underestimates the expected deaths in 2020 under the counterfactual no-COVID scenario, thereby biasing excess deaths.

## Data and Methods

### Raw and derived data

To replicate the more elaborate expected death regression models, we collected cross-country data on weekly death counts and population exposure by age and sex, along with information on the timing of public holidays and weekly population-weighted temperature anomalies (Figure S.2).

Age and sex-specific weekly death counts by country were sourced from the Short-term Mortality Fluctuations (STMF) database [13, 21]. We selected all European countries with data going back to at least 2007. As we are using the Mean Absolute Percentage Error (MAPE) measure to report on prediction error, we needed to ensure strictly positive weekly death counts. We thus excluded Estonia, Iceland, and Luxembourg from the analysis and summed deaths into age categories 0 to 65, 65 to 75, 75 to 85, and 85+.

Person-weeks of population exposure by country, sex, and age were derived from mid-year population estimates from the Human Mortality Database [11]. We summed these estimates into the target age groups and then interpolated over the weeks of a year by fitting a cubic spline and extrapolating linearly to the end of 2020. Integrating the spline over single weeks yielded person-weeks of exposure.

Population weighted weekly temperature anomalies were calculated from global gridded temperature [7] and population data [6]. First, we calculated the average weekly temperature per grid cell from the daily measurements and then weighted these measurements by the population size in the same grid cell. We then selected all grid-cells overlapping with the area of a given country and calculated the country’s population-weighted average weekly temperature. By averaging the weekly temperatures over multiple years, we established an expected temperature. A country’s weekly population-weighted temperature anomalies were then derived by subtracting this long-term trend from the weekly temperature in a given year.

We used the “Nager.Date” software [10] to determine for each week, year, and country the occurrence and type of a public holiday, distinguishing between Christmas, Easter, New Year, and “other” types of public holidays.

### Model choice

Expected deaths and associated excess deaths statistics were computed under nine different models listed in Table 1. The models were chosen to represent the range of approaches in the literature on excess deaths estimation since the start of the pandemic, from very simple to elaborate. All models predict age and sex specific expected death counts and are fitted separately for each of the 20 countries.

With the exception of the Latent Gaussian Models (LGMr, LGMrt) all models were specified as quasi-Poisson regressions on weekly death counts *D*_*i*_ of the form

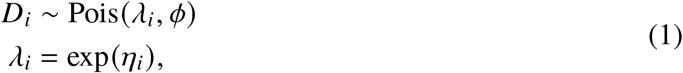

where *λ*_*i*_ is the expected death count, *ϕ* is the dispersion factor, and *η*_*i*_ is the linear predictor on a log-scale. While we considered employing the negative binomial distribution instead, convergence issues due to sporadic underdispersion informed our choice of the more robust quasi-Poisson model.

For the *5 year average death count model* (AVGc5) we specified

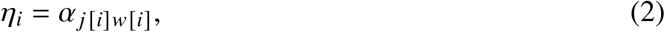

where *α*_*j*[*i*] *w*[*i*]_ is a coefficient for iso-week *w* and sex × age stratum *j* of observation *i* within some country. The model was fit over a period of five years.

Log-person-weeks exposure log *E*_*i*_ were added as an offset to (2), yielding the *5 year average death rate model*

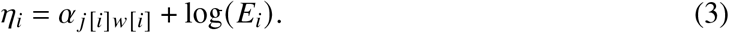

The *Serfling with exposures* (SRFr) model was specified as

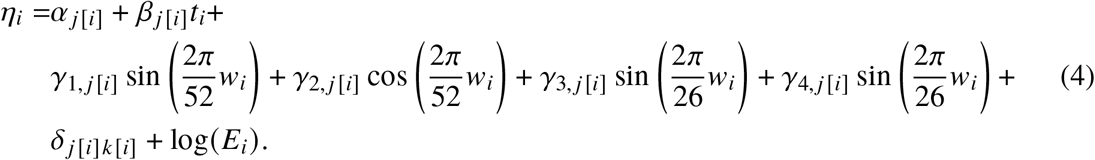

Model (4) features log-linear time trends *β*_*j*[*i*]_ over weeks *t*_*i*_ since the earliest observation, parametric seasonal effects *γ*_*j*[*i*]_, and the effect of a public holiday δ_*j*[*i*]*k*[*i*]_ on the expected death counts, where *k* denotes the type of holiday. The model is completely stratified over sex and age groups *j*. Within each stratum we selected the number of seasonal terms (no seasonality or *γ*_1,2_ or *γ*_1,2,3,4_) via AIC using the likelihood from an initial Poisson regression without overdispersion. Omitting the term log *E*_*i*_ from (4) yields the *Serfling without exposures* (SRFc) model. For the *Euromomo style Serfling model* (SRFcem) we dropped the exposure as well and only fitted the model on weeks 15 through 26 and 36 through 45. All Serfling models were fitted via the `glm` function in `R`.

The *Generalized Additive Model with temperature anomaly* (GAMrt) allows for non-parametric, smooth seasonal effects and adjusts for the effect of extreme temperature on weekly death counts. We specified the model as

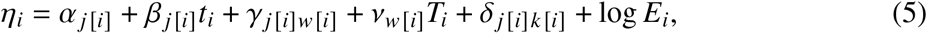

where *γ*_*j*[*i*]*w*[*i*]_ = *f*_*j*[*i*]_ (*w*_*i*_) is a stratum specific seasonality term implemented as a cyclical penalized spline with 12 knots spaced in equal distance over the weeks of a year *w*_*i*_, and *v*_*w*[*i*]_ = *g* (*w*_*i*_) is the temperature anomaly coefficient varying smoothly over *w*_*i*_ in a cyclical fashion. For the *Generalized Additive Model without temperature anomaly* (GAMr) we omitted this term. Analogous to the model (4) we specified a log-linear trend, public holiday intercepts, and an offset controlling for population exposure. The model was fitted via the `gam` function from the `mgcv` library in `R`.

The *Latent Gaussian Model with temperature anomaly* (LGMrt) follows the specification by [14]. It features a nonparametric seasonal effect which may vary over time and an autoregressive trend. The model is fitted separately by age group and sex and, omitting the subscript *j*[*i*], can be written as

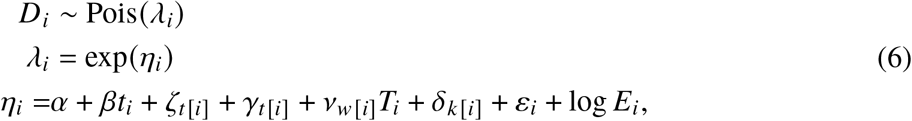

where *βt*_*i*_ is a simple log-linear trend over time and 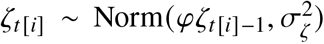 is a first order autoregressive trend. In contrast with the other models over-dispersion is directly included in the linear predictor in form of the unstructured error term 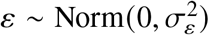. Seasonality is captured with a time-varying random coefficient *γ*_*t*[*i*]_, following [23] section 3.5, where the summed effect of every consecutive series of 52 weeks on mortality is distributed as 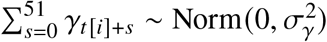. The effect of temperature anomalies on weekly mortality is captured by the varying coefficient 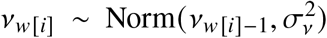, specified as a circular first order random walk over week of year. The temperature effect was dropped for the *Latent Gaussian Model without temperature anomaly* (LGMr). The models were fitted with the `INLA` library [24, 25] in `R`.

### Excess death measures

We defined excess deaths *D*^∗^ in a given week *t* and stratum *j* as 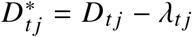, where *D* is the observed death count and *λ* are the expected deaths under a given model. This definition as a model residual allows to capture mortality displacement effects where a positive excess in part of the year can be followed by negative excess in later weeks. The total number of excess deaths over period [*a,b*] are given by 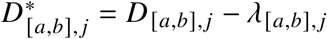, where 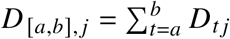 and 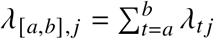. Following a common strategy, expected and observed deaths were summed over strata to estimate the excess at a higher aggregation level [3, 1, 14, 31].

To facilitate comparisons between countries, we report the weekly percentage increase of observed deaths over expected deaths, 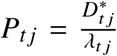, the so-called “P-score”. For a sequence of weeks [*a, b*] we calculated the cumulative P-score from the total excess and expected deaths over that period,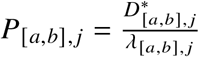.

### Prediction intervals

Prediction intervals around the measures of excess death were computed via simulation. We first sampled 500 sets of parameter estimates, *s* = {1, …, 500}, either from a multivariate normal distribution Norm (*β*, ∑), with *β* being the vector of estimated parameters, and ∑ the variance-covariance matrix of the fitted model, or, in case of the Latent Gaussian Models, from the posterior distribution of the parameters. For each of the 500 simulated sets of parameters we calculated the expected weekly death count 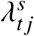 and then sampled a single simulated death count 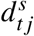 from a negative-binomial distribution with mean equal to 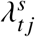, and variance equal to 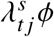, matching the characteristic moments of the quasi-Poisson distribution. In case of under-dispersion, where *ϕ <* 1, and for the Latent Gaussian Models, we sampled from a Poisson distribution with rate 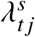. The simulated death counts 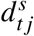 were then substituted into the excess death formulas in place of the expected deaths *λ*_*tj*_ and the resulting distribution of excess summarized by its quantiles.

### Cross-validation

In order to assess the accuracy of the predicted death counts from each model we performed a cross-validation analysis. Time series of weekly death counts and co-variates were split into five cross-validation sets (Figure S.1). The training period starts at week 27 of a year and ends after seven fully observed summer to summer seasonal cycles with the beginning of week 8 of a year. After fitting the model, weekly death counts were predicted for 45 weeks following week 8 of the last year in the training set. The starting years for the five cross-validation series were 2007 to 2011. This particular cross-validation setup has been chosen to mirror the task of predicting weekly deaths in 2020, given data observed until shortly before the outbreak of the pandemic.

We compared the expected death counts *λ*_*i*_ with the observed death counts *D*_*i*_ in the test set and quantified the prediction bias via the mean percentage error, 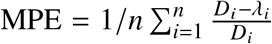, and the prediction error via the mean absolute percentage error, 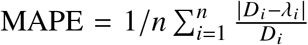, where *i* runs over the *n* predictions in the five test sets.

All predictions were made by country, week, sex, and age. To facilitate presentation we first summed the observed and predicted death counts a) over weeks, sex, and age (annual deaths by country), and b) over sex and age alone (weekly deaths by country), then calculated the aggregated prediction bias and error by country, and finally summarized the distribution of these aggregated measures of model fit across countries via quantiles. Stratum-specific prediction errors can be found in the appendix.

## Results

Models agreed on the weekly *pattern* of excess deaths but differed in the estimated *level* of excess (Figure 1). The following weekly *patterns* were identified by all models: Austria, Switzerland and the Eastern European countries suffered higher weekly peak excess deaths in fall and winter than in spring. The situation was reversed in France, Netherlands, Scotland, Spain, and Sweden, with lower levels of weekly percent excess in the second half of the year. Belgium and Portugal experienced three distinct waves of pronounced excess death in 2020. Disagreement among models regarding the *level* of country specific weekly and annual percent excess deaths in 2020 was typically around 5%, but could reach double digits within some age and sex strata (Figures S.3–S.10). Consequently, models disagreed at times regarding the existence of significantly elevated annual percent excess (*P*_2020_ *>* 0 with probability ≥ 0.95). On the country level we found such disagreement only in Denmark, Finland, Latvia, and Norway, countries with relatively few registered COVID deaths. Within age and sex strata model disagreement was more widespread, affecting 15 out of 20 countries. Belgium, Spain, Scotland, Portugal, and Slovenia were the only countries where all nine models agreed, within *all* age and sex strata, that annual percent excess deaths have been significantly elevated above zero in 2020. On the country level the average death rate model (AVGr5) produced the lowest annual percent excess death estimates in 18 out of 20 countries, whereas the maximum varied, with the Euromomo style Serfling (SRFcem) and the average death count model (AVGc5) producing the highest excess in 13 out of 20 countries.

**Figure 1:**
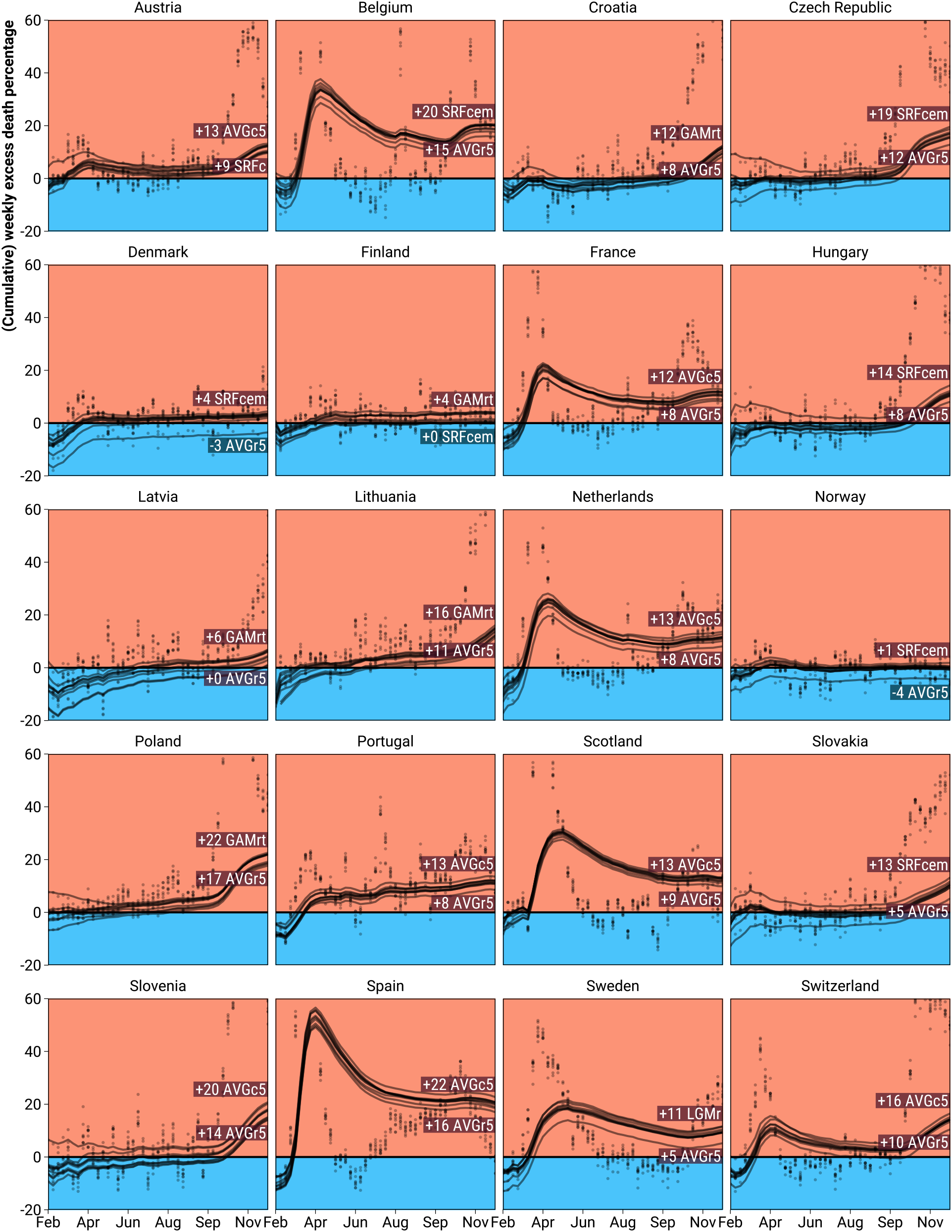
Weekly and cumulative excess death percentage as predicted from 9 different models during the year 2020 weeks 8 through 52 for 20 European regions. *Note: Points indicate weekly excess percentages. Cumulative excess percentages are indicated by curves and derived from cumulated observed and expected deaths. The labels refer to the range of predicted percent excess over the entire analysis period. Y-axis truncated at 60%*.

The ranking of countries along the percent excess deaths in 2020 changed between models (Figure 2). All models agreed on the five countries with the highest annual percent excess (Poland, Spain, Belgium, Slovenia, Czech Republic) and on the four least affected countries (Finland, Latvia, Denmark, Norway). There was substantial disagreement regarding the middle ranks with Hungary, Lithuania, Sweden, Slovakia, Austria, Hungary, Netherlands, and Portugal jumping by more than 5 positions between models. Ranks under the Serfling with exposures (SRFr) and Generalized Additive Models (GAMr, GAMrt) exhibited high agreement. Within age and sex strata we observed more drastic changes in rank across models, but again with good agreement concerning the top and bottom ranks (Figure S.11).

**Figure 2:**
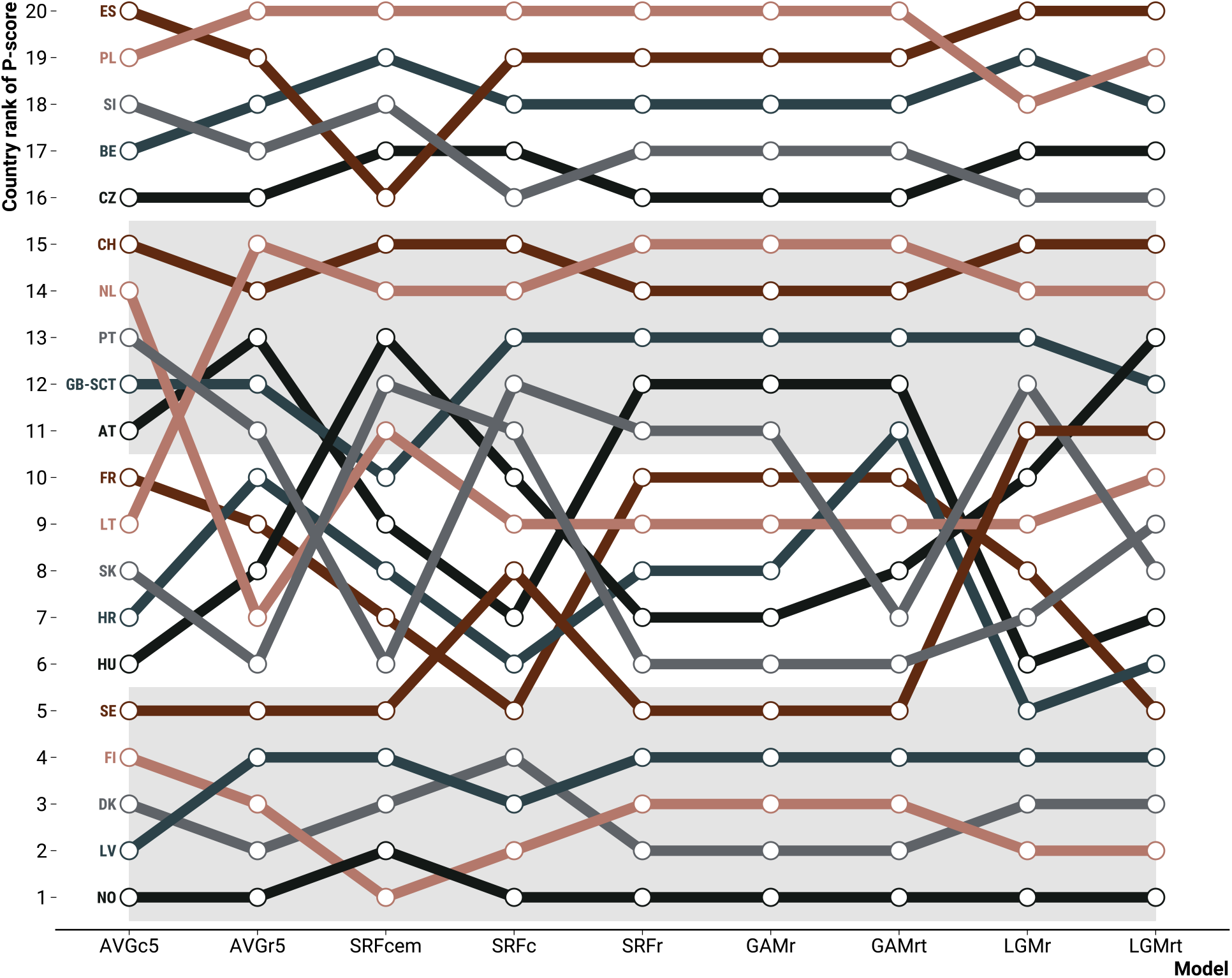
Country ranking of excess death percentage during the year 2020 weeks 8 through 52 under 9 different models.

We compared the 5-year average death count model (AVGc5, the reference) to various other models concerning the expected number of deaths in 2020 (Figure 3). With the exception of the Euromomo Serfling model (SRFcem), all models tended to predict higher numbers of weekly and annual deaths compared to the average death count (AVGc5) model. The 5-year average death rate model (AVGr5) gravitated towards the highest expected death counts, with annual predictions being higher by 3.1 to 5.1% compared to the reference for half of the countries. We also found rather high baseline estimates for the Latent Gaussian Models (LGMr, LGMrt) and the Serfling without exposures (SRFc) model. Within population strata results varied. The tendency for the AVGc5 model to produce low expected death counts was most pronounced in the age group 85+, but clearly reversed in the age group 0 to 65. Likewise, the tendency for the AVGr5 model to produce high estimates of excess was present in all age groups but the highest (S.12).

**Figure 3:**
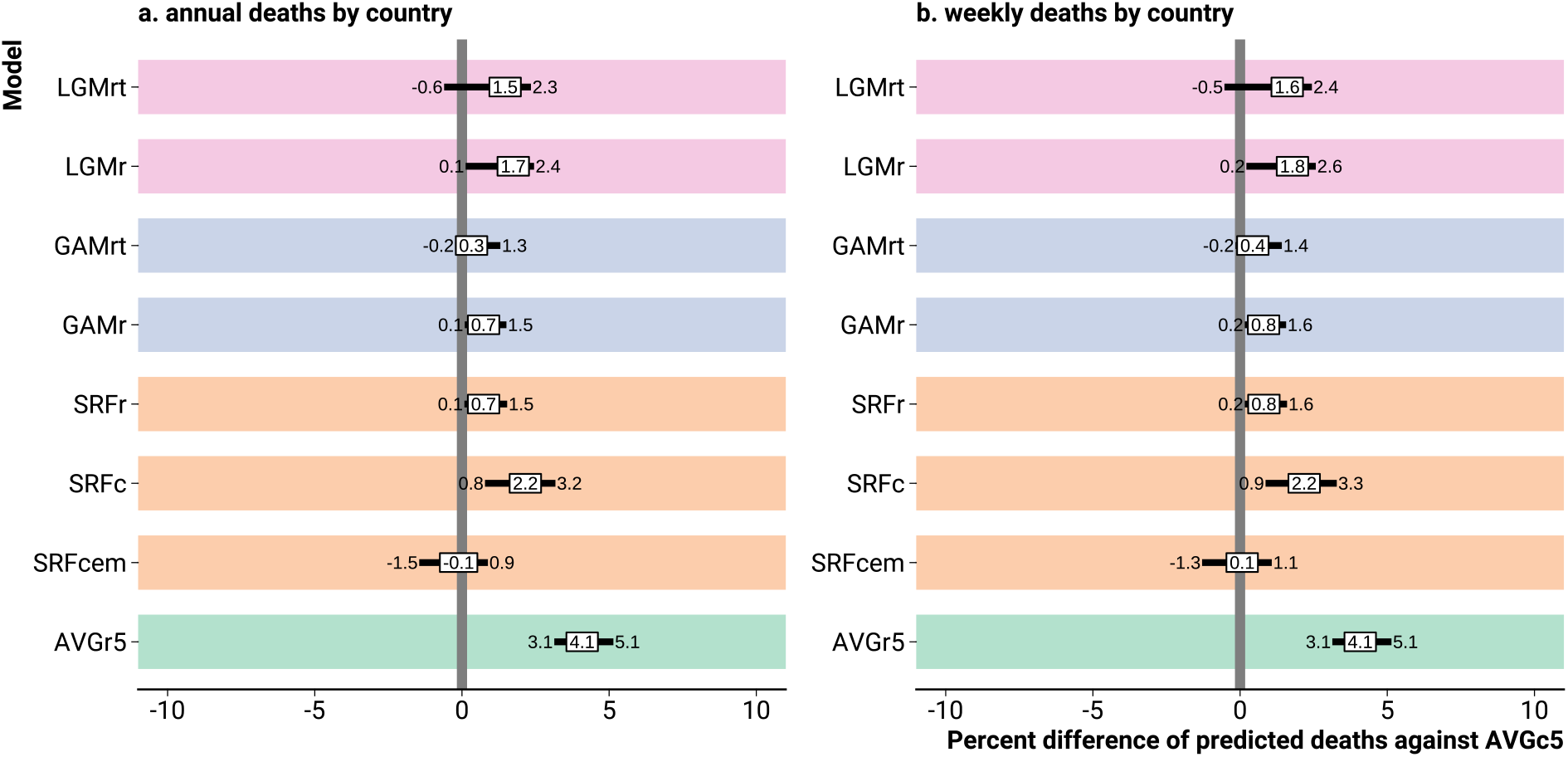
Percent differences of predicted death counts from various models against the 5-year average weekly death count model prediction. *Note: The prediction differences were summarised across countries by the 0*.*25, 0*.*5, and 0*.*75 quantiles*.

The cross-validation study confirmed the substantial biases of the Serfling Euromomo regression and the 5-year average death rate and count models (Figure 4). While the average rate model (AVGr5) consistently displayed the highest propensity among all models to overestimate annual or weekly deaths on the country level, the average count (AVGc5) and the Serfling Euromomo models (SRFcem) underestimated deaths. The lowest bias was presented by the models GAMr and SRFr. For both models the average percentage error when predicting annual death counts was between − 1% and 0.2% for half of the countries with similar results for week-by-week predictions. Omitting the population structure term from a Serfling model increased the prediction bias as seen for model SRFc. Between sex and age strata the relative model performance when predicting annual deaths often varied with model AVGc5 changing direction of the bias over age and model SRFcem being comparably unbiased in the age group 85+ (Figure S.13). Models GAMr and SRFr consistently displayed low bias across strata. We found no evidence that the inclusion of a temperature variable decreased the prediction bias. The Latent Gaussian models (LGMr, LGMrt) exhibited a tendency for overprediction, but less severe than seen with model AVGr5.

**Figure 4:**
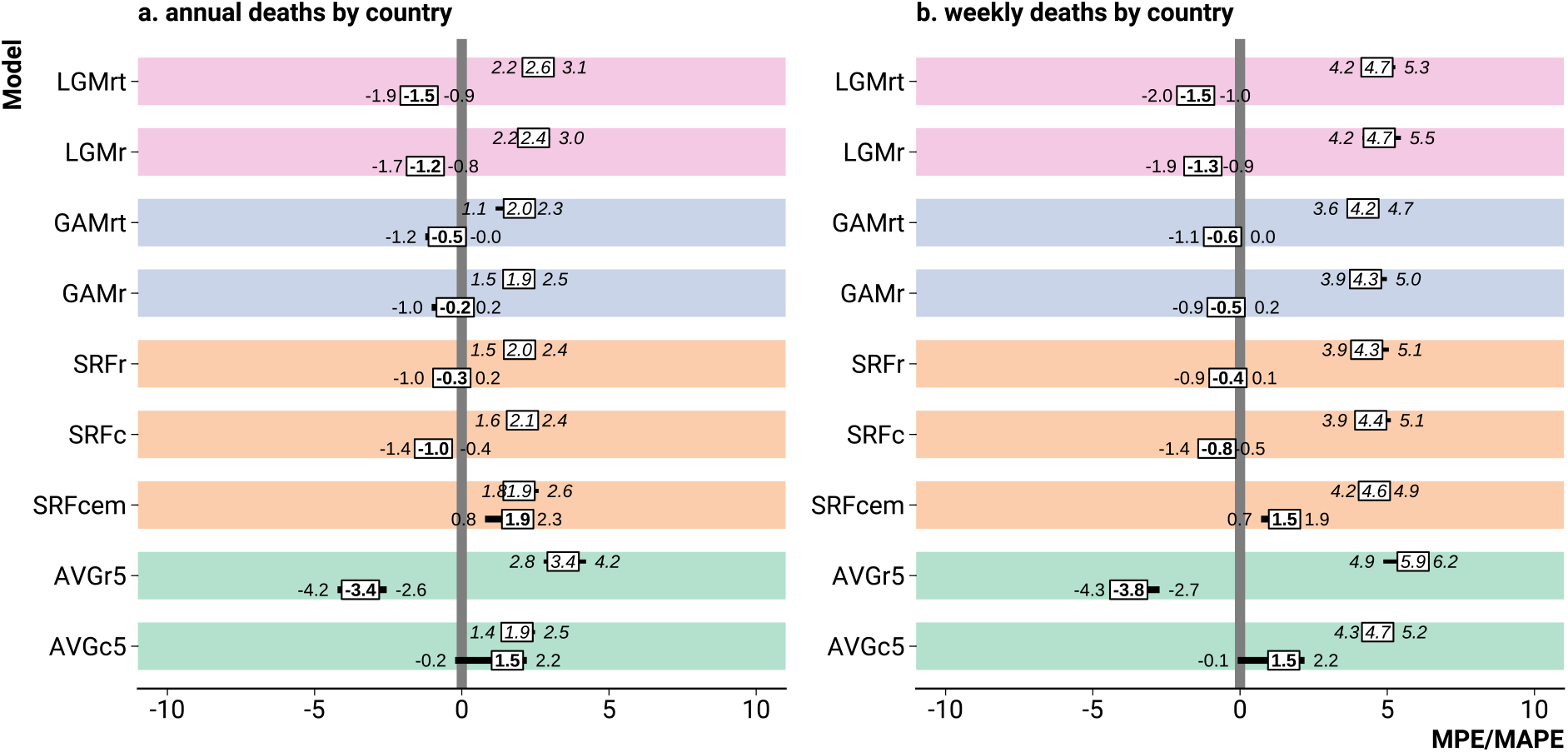
Bias (MPE, bold) and error (MAPE, italic) by model when predicting death counts on test data. *Note: The error and bias measures were summarised across countries by the 0*.*25, 0*.*5, and 0*.*75 quantiles*.

Prediction errors tended to more than double going from annual to weekly predictions and varied across countries (Figure 4). Annual death counts were predicted with a MAPE ranging from 1.9–3.4% across models, increasing to 4.2–5.9% for the weekly predictions. Notably, the annual country level predictions from the simple AVGc and the biased SRFcem model exhibited low MAPEs. This result however, did not extend to annual predictions by age and sex, where both models showed varying biases and comparatively high errors (Figure S.13). Because the annual predictions were aggregated from the stratum specific predictions, this suggests a potential for errors to cancel out in the aggregation process. Within strata the GAM and SRFr models exhibited the lowest error when predicting annual deaths and the relative disadvantage of the LGM models increased with age. Omitting the exposure variable from the Serfling model increased the MAPE. The inclusion of a temperature effect did not consistenly decrease the prediction error.

## Discussion

No matter the chosen model, in 2020, deaths across Europe have been unusually elevated, exhibiting a weekly pattern closely following the course of the COVID-19 pandemic in the different countries. Other inferences, however, are less robust to the choice of model. Depending on how the expected deaths were calculated, one may have either Poland or Spain as the worst affected country, Denmark may or may not have had significantly elevated death counts, annual deaths among Slovakian males ages 85+ may be increased by as little as 4% to as much as 15%, and death counts following the first wave of the pandemic may have returned to expected levels or dropped into a “mortality deficit.” Thus, short of a standard procedure of specifying expected deaths, robustness checks are crucial. In the simplest case, such a check could be based upon expected deaths via 5-year average weekly death rates and alternatively a Serfling model similar to the Euromomo implementation, as we found these two approaches to have opposite biases, with predictions from other methods often falling in-between.

The Euromomo model is designed to predict death counts as expected under exceptionally mild seasonal mortality increases, thus resulting in high estimates of excess death. While this makes the Euromomo approach very useful for monitoring influenza-related mortality, the baseline is biased if interpreted as “expected deaths in the absence of the COVID-19 pandemic”, as the typical seasonal increase in deaths is not as shallow as predicted under Euromomo. The technical reason for the low Euromomo predictions during fall and winter is the exclusion of weeks with likely influenza activity from the model fitting. If the modeling goal is mortality forecasting, as under the counterfactual no-COVID scenario, then fitting seasonal regression models over all weeks of a year reduces the bias in the forecasted death counts.

A lack of adjustment for continuous mortality improvements, as have been observed across Europe up until 2020 [15, 2], explains the tendency of the average death rate model towards low estimates of excess. By simply averaging declining death rates over time, one overestimates expected mortality for the current year, thus underestimating excess. Claims of no excess deaths based upon averaged weekly death rates should therefore be checked for robustness against the inclusion of a time trend.

Two opposing time-trends explain the bias of the ubiquitous average weekly death count model: increasing deaths due to population aging and decreasing deaths due to mortality improvements. As the model adjusts for neither trend, it may be biased in either direction, depending on the relative size of either effect in any given country, although we found a slight tendency towards high excess death estimates across Europe. The lower overall bias when averaging counts instead of rates suggests that errors due to non-adjusted time trends cancel to some degree, demonstrating that an incomplete adjustment to trends can be worse than no adjustment at all.

A major challenge faced by every model is to estimate the magnitude of the future seasonality. In the absence of information about influenza activity, all models just predicted an average seasonality for the following Fall and Winter, possibly with minor temperature adjustments. Whether or not an influenza predictor should be included when estimating excess deaths over the COVID-19 pandemic depends on the purpose of the analysis: causal inference or mortality monitoring. A causal estimate of the direct and indirect effects of the pandemic on overall mortality can not accommodate observed influenza activity past the beginning of the pandemic as this variable is itself influenced by the “treatment,” i.e., COVID-19 and associated mitigation measures [8, 28]. In a counterfactual world where the COVID-19 pandemic did not happen, influenza activity throughout 2020 may have been different. For mortality monitoring purposes, however, the inclusion of an influenza predictor is admissible as the question is whether the currently observed death counts are unusual given the currently observed levels of influenza.

How many deaths would we have expected in 2020 if this would have been a typical year? This definition of expected deaths allows to discriminate between suitable and unsuitable models on empirical grounds: We want a model that excels at short-term forecast of weekly death counts. While robustness checks and model averaging are pragmatic way to deal with model specification uncertainty, models of expected deaths are not beyond scrutiny – like other prediction models, they can and should be put to the test. Further research into “short-term mortality prediction” promises a better understanding of systematic biases across different model specifications, which can then be synthesized into a set of best practices when modeling expected deaths in the absence of unusual events.

## Data Availability

Data and code to replicate the results in this paper are available at https://github.com/jschoeley/rbx2020.

## Supplementary materials

### Supplementary figures

**Figure S.1:**
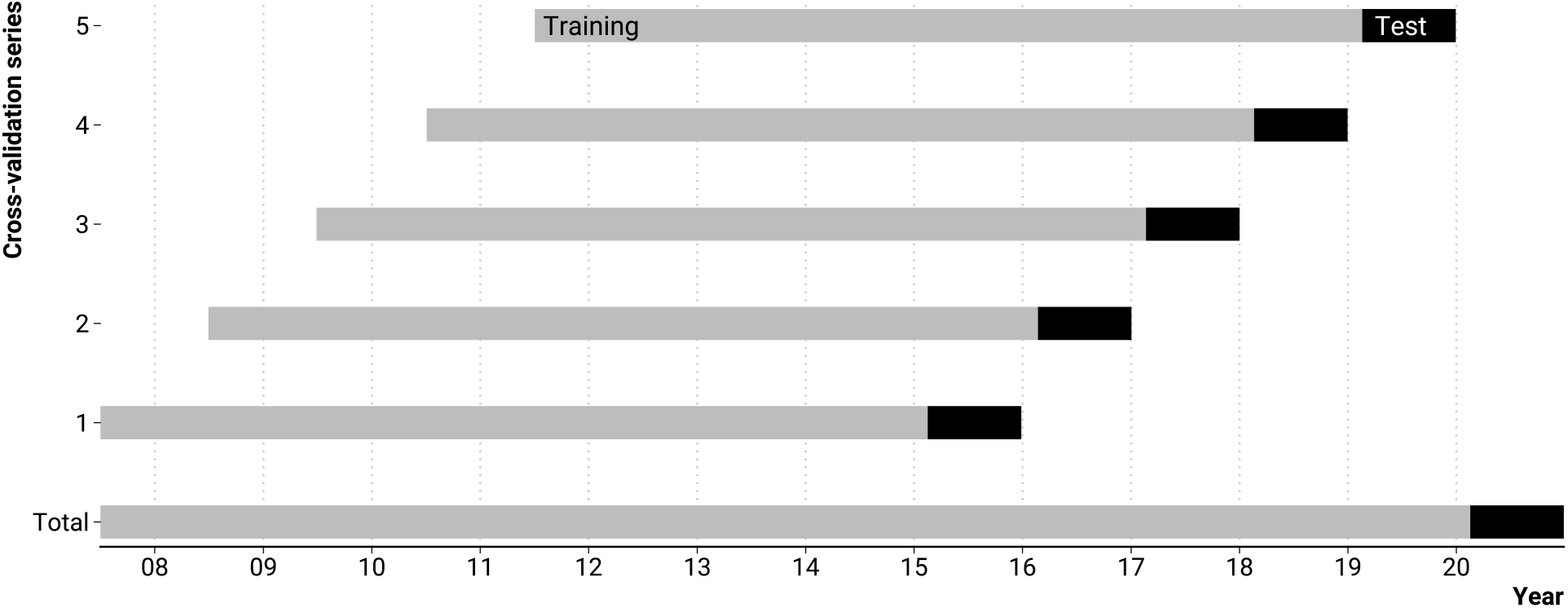
Rolling origin five-fold cross-validation setup mirroring the task of predicting weekly deaths past the beginning of the COVID pandemic given pre-pandemic data.

**Figure S.2:**
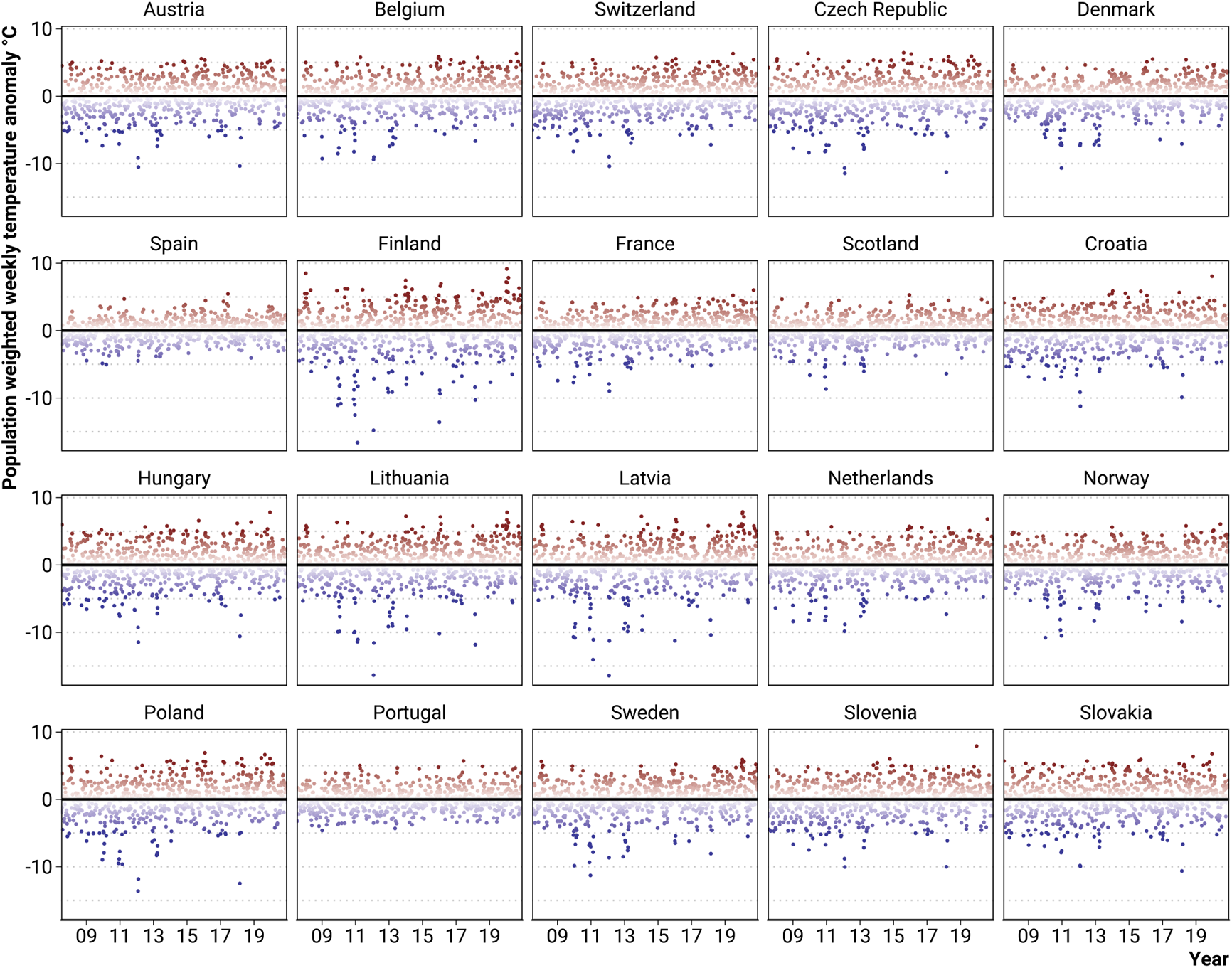
Population weighted weekly temperature anomaly by country.

**Figure S.3:**
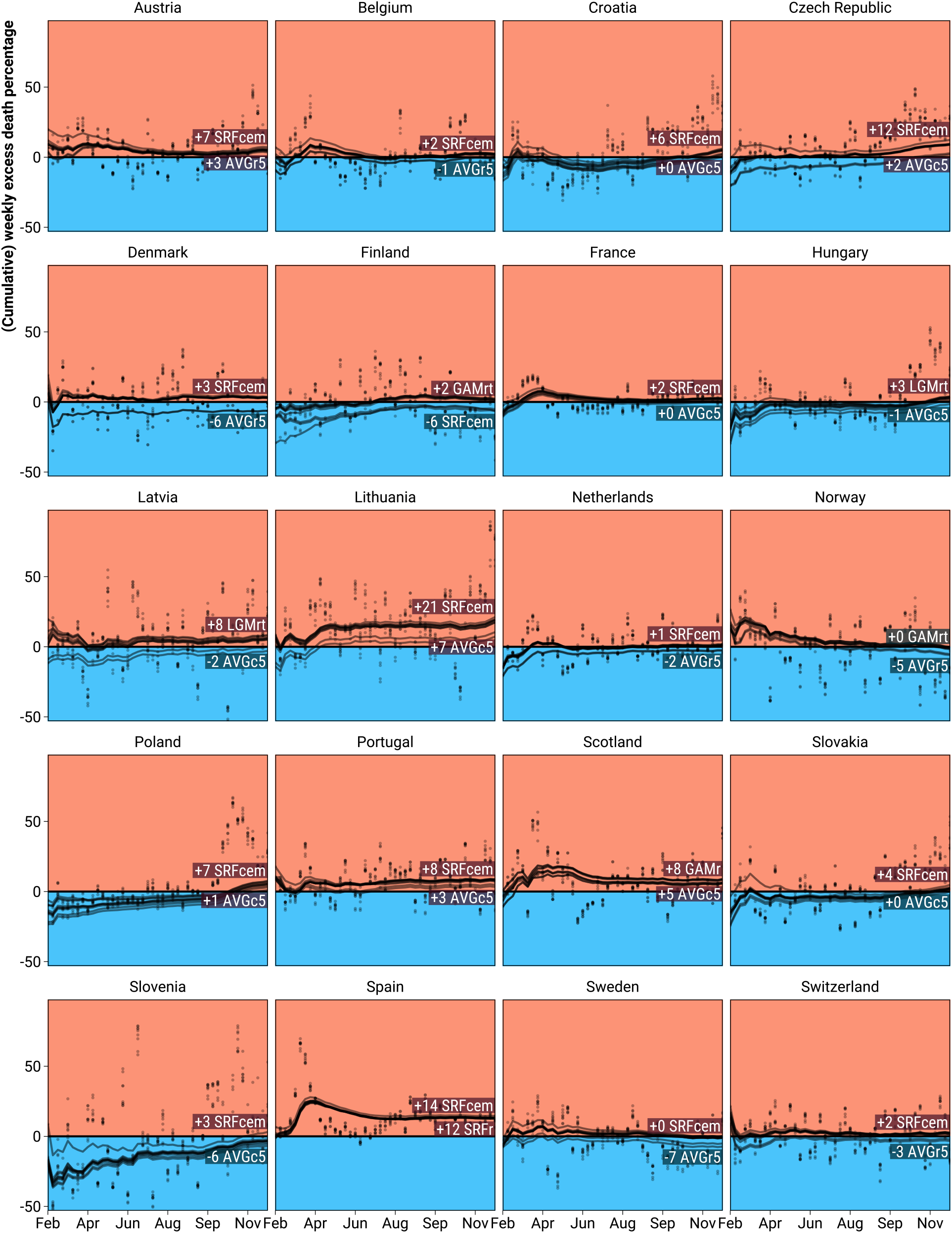
Female percent excess deaths for ages 0 to 65 as predicted from 9 different models during the year 2020 weeks 8 through 52 for 20 European regions.

**Figure S.4:**
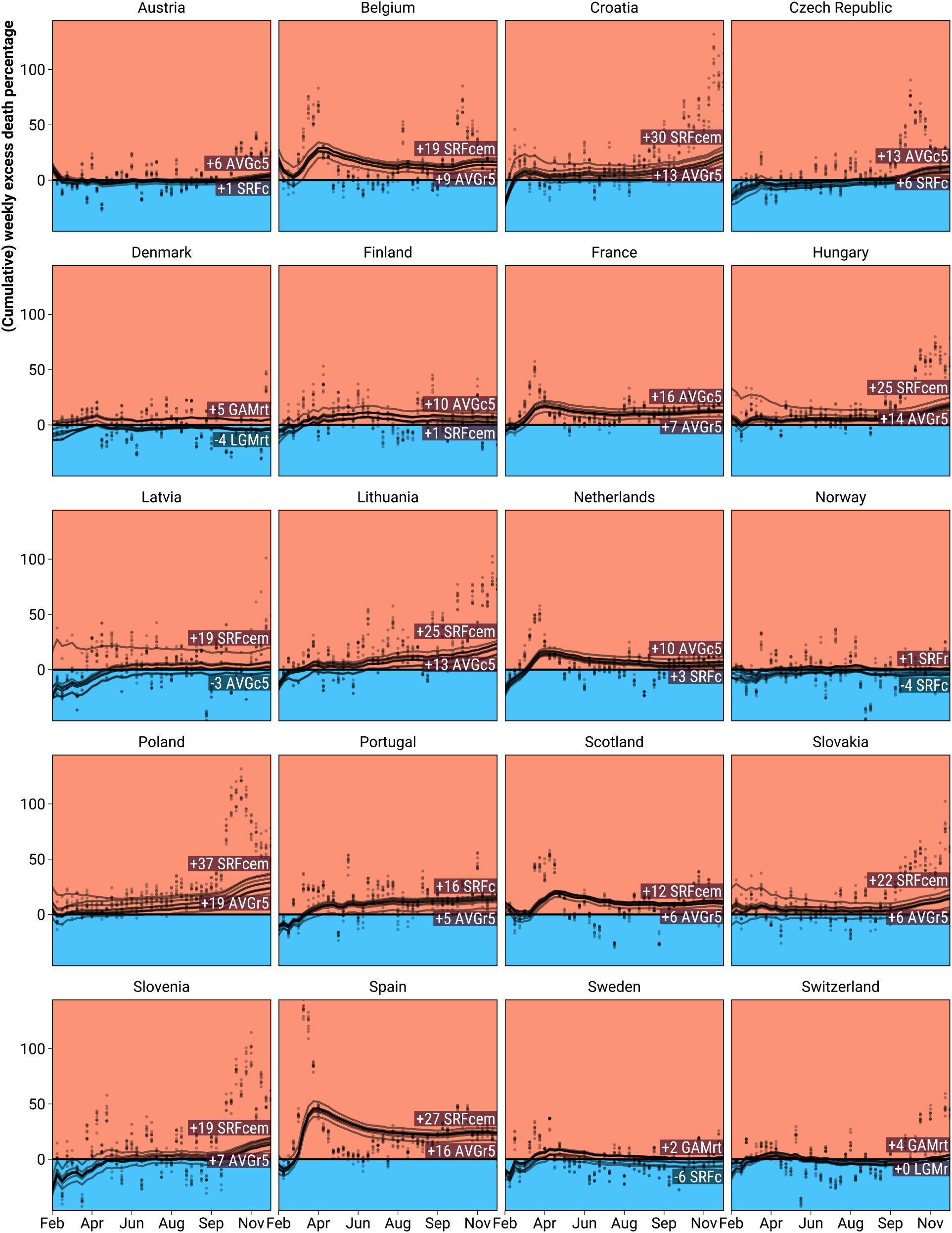
Female percent excess deaths for ages 65 to 75 as predicted from 9 different models during the year 2020 weeks 8 through 52 for 20 European regions.

**Figure S.5:**
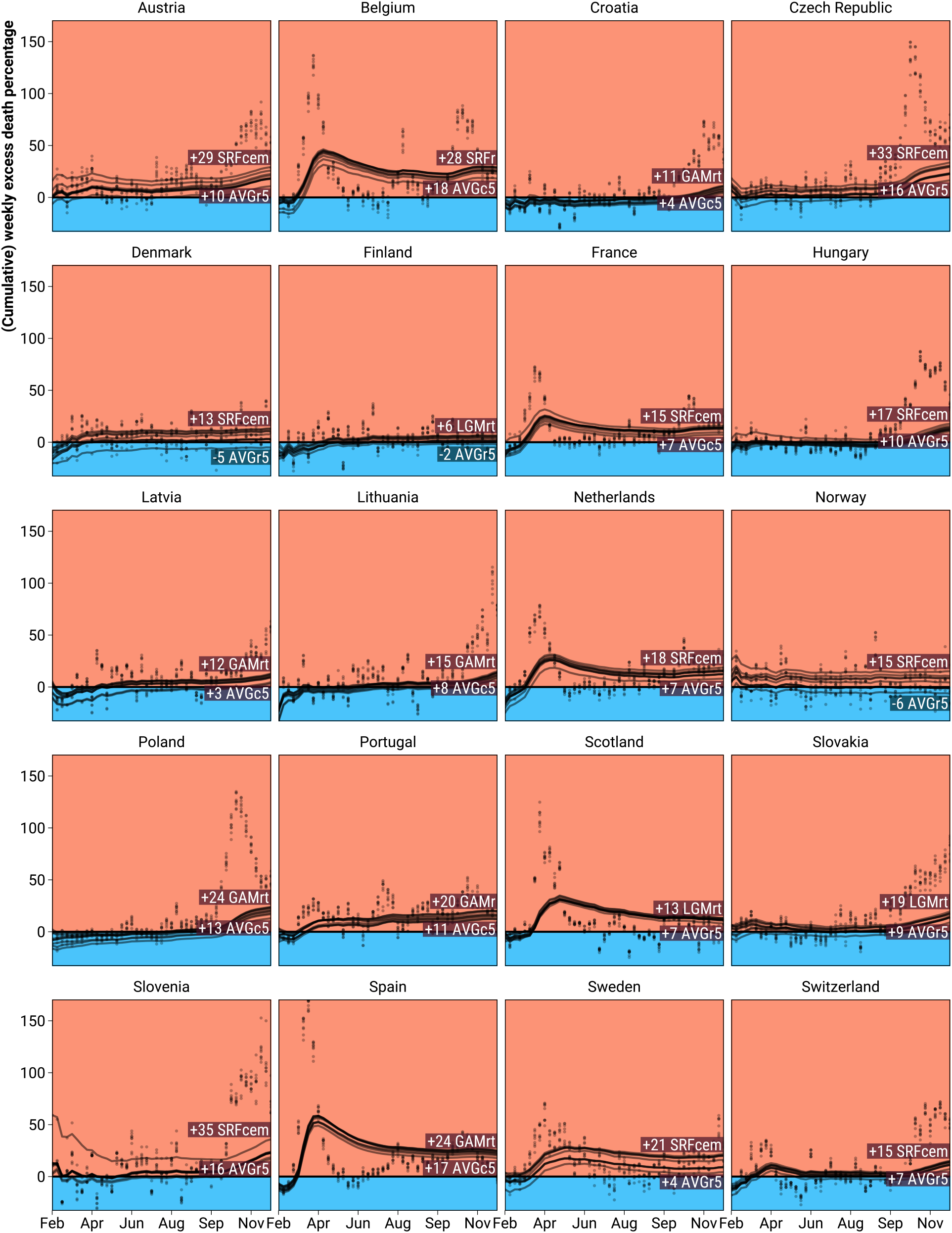
Female percent excess deaths for ages 75 to 85 as predicted from 9 different models during the year 2020 weeks 8 through 52 for 20 European regions.

**Figure S.6:**
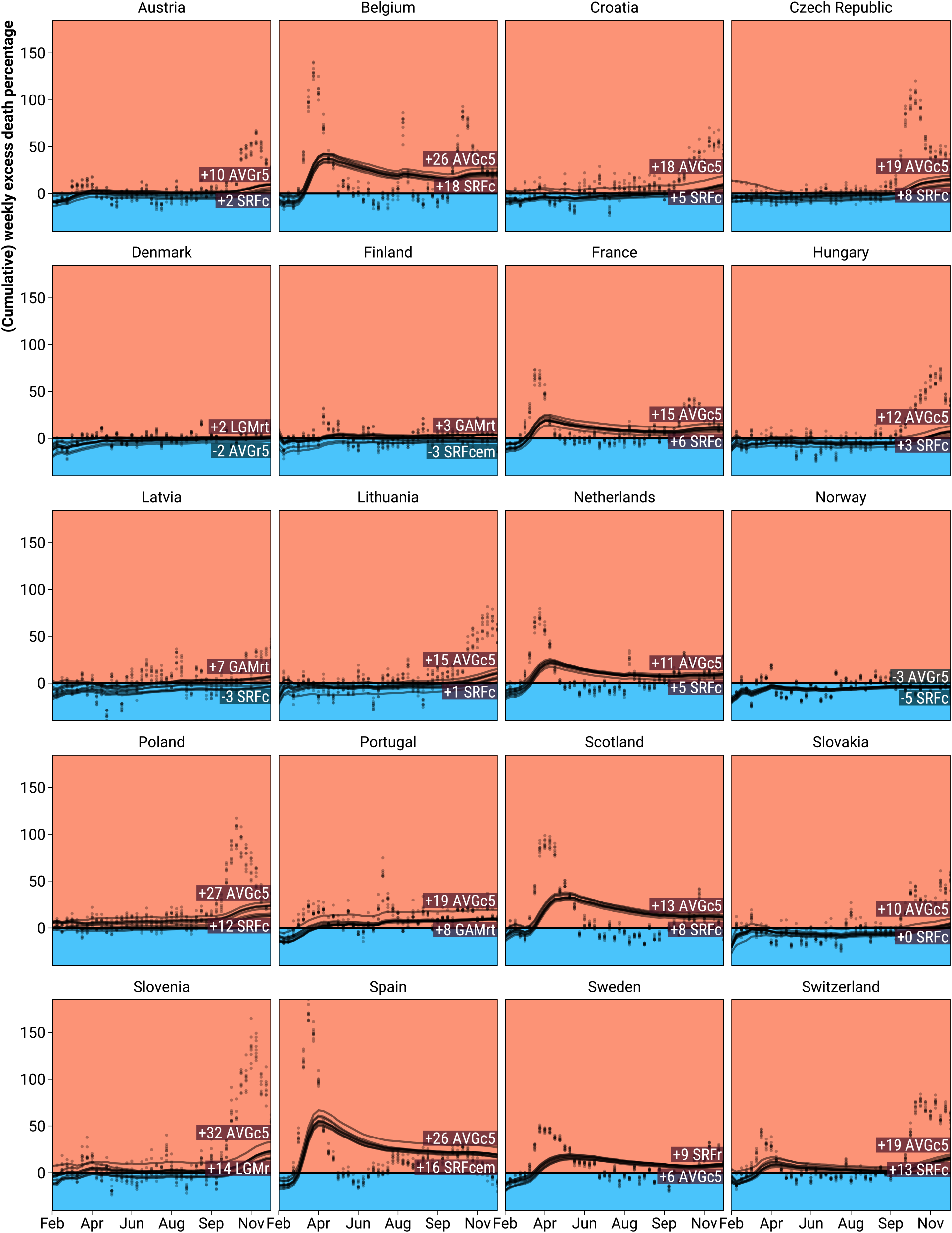
Female percent excess deaths for ages 85+ as predicted from 9 different models during the year 2020 weeks 8 through 52 for 20 European regions.

**Figure S.7:**
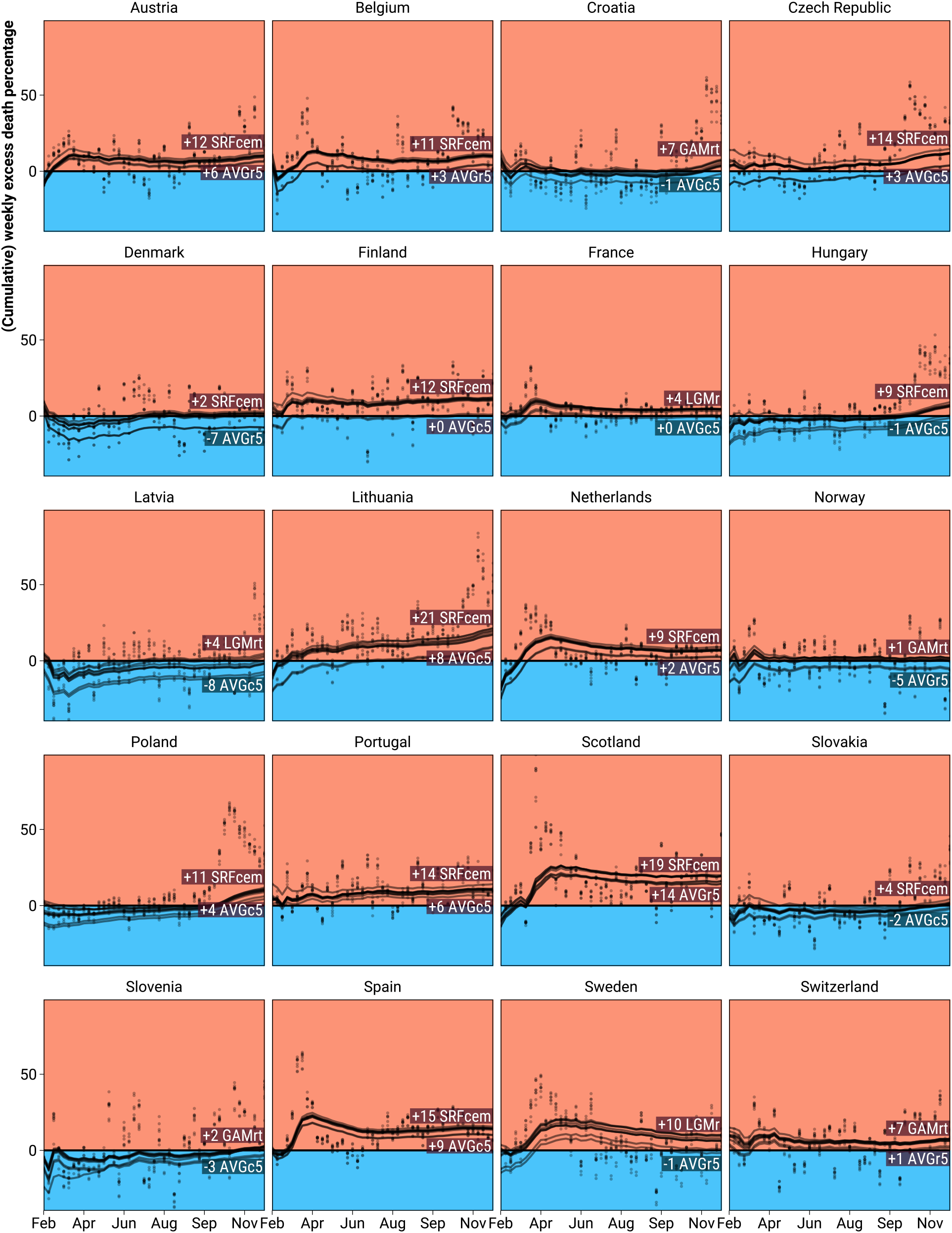
Male percent excess deaths for ages 0 to 65 as predicted from 9 different models during the year 2020 weeks 8 through 52 for 20 European regions.

**Figure S.8:**
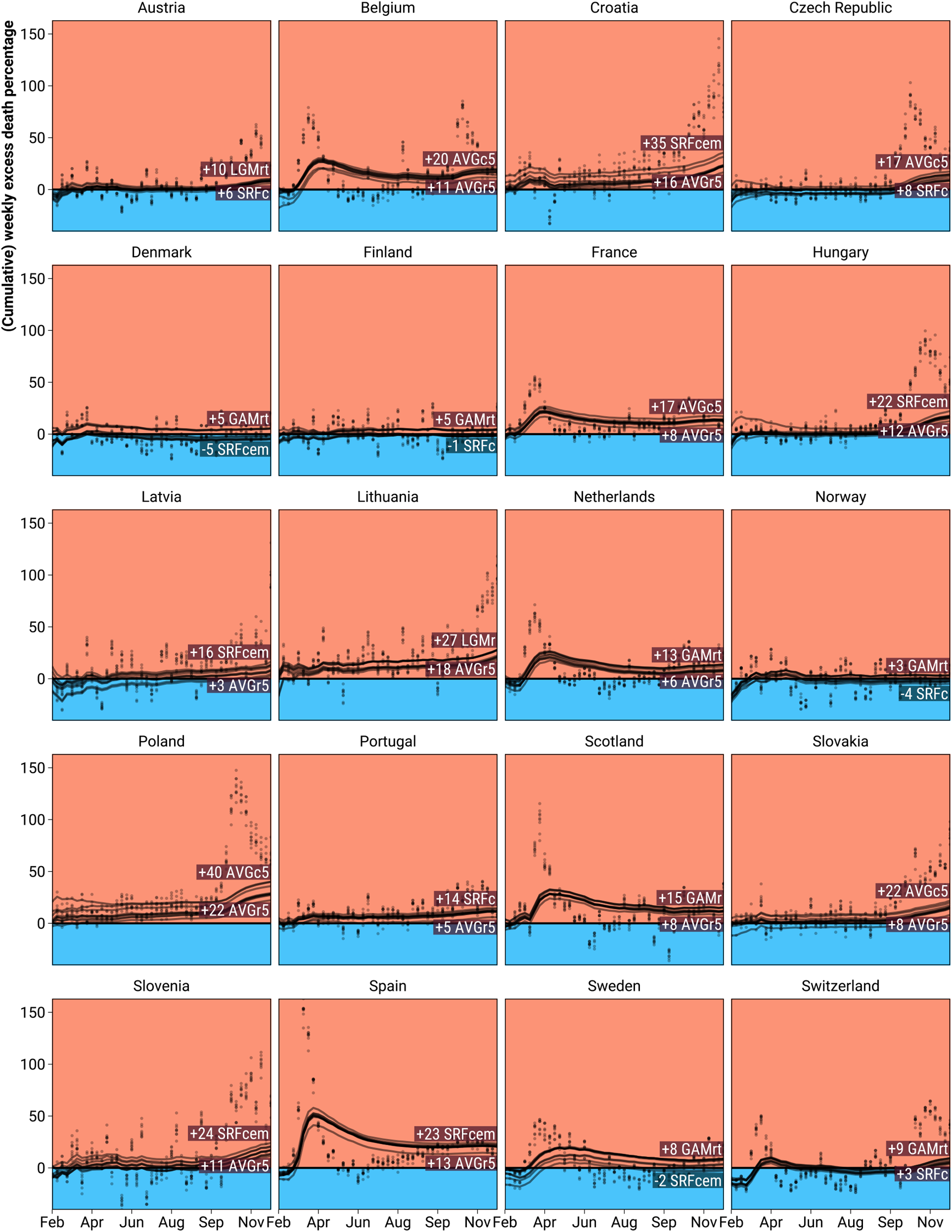
Male percent excess deaths for ages 65 to 75 as predicted from 9 different models during the year 2020 weeks 8 through 52 for 20 European regions.

**Figure S.9:**
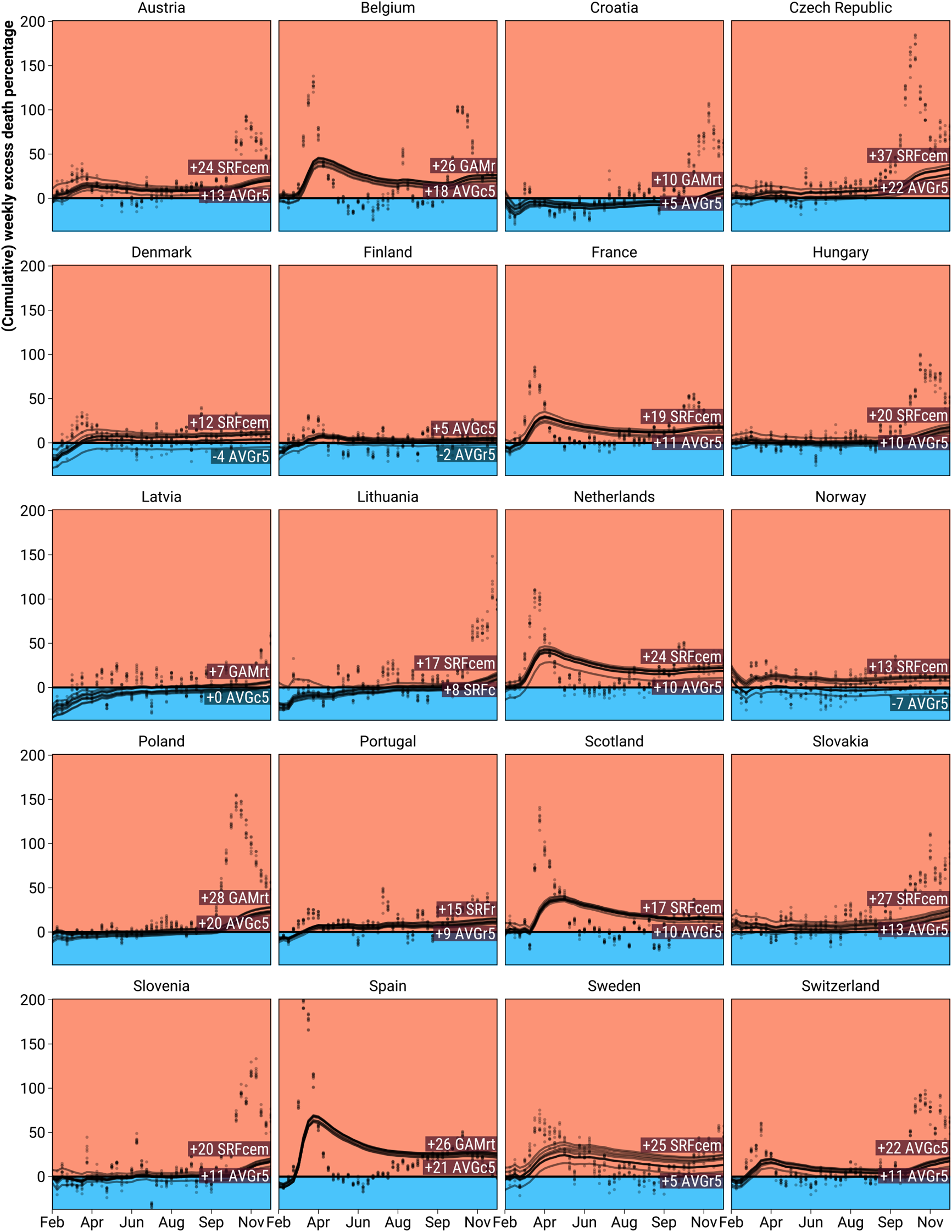
Male percent excess deaths for ages 75 to 85 as predicted from 9 different models during the year 2020 weeks 8 through 52 for 20 European regions.

**Figure S.10:**
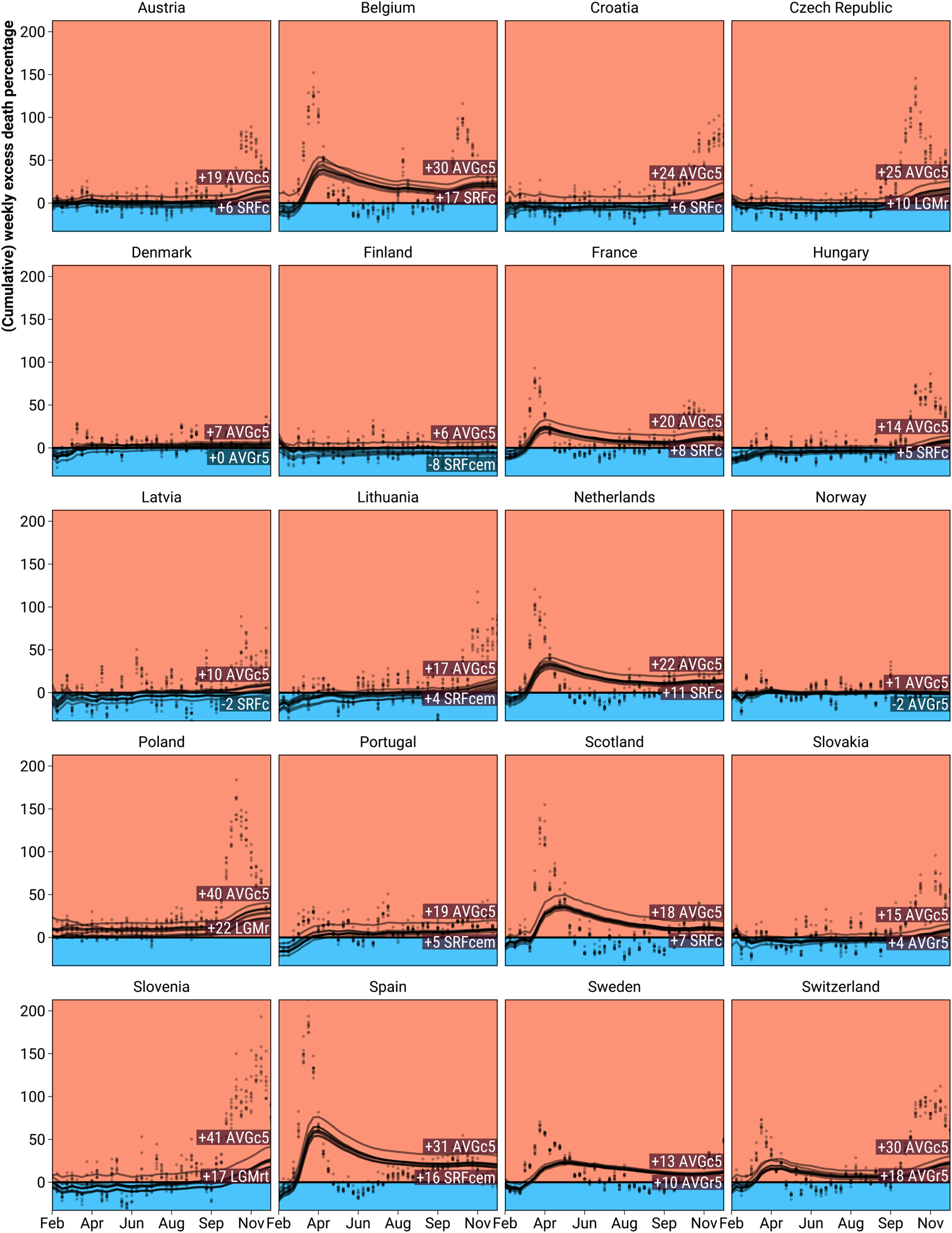
Male percent excess deaths for ages 85+ as predicted from 9 different models during the year 2020 weeks 8 through 52 for 20 European regions.

**Figure S.11:**
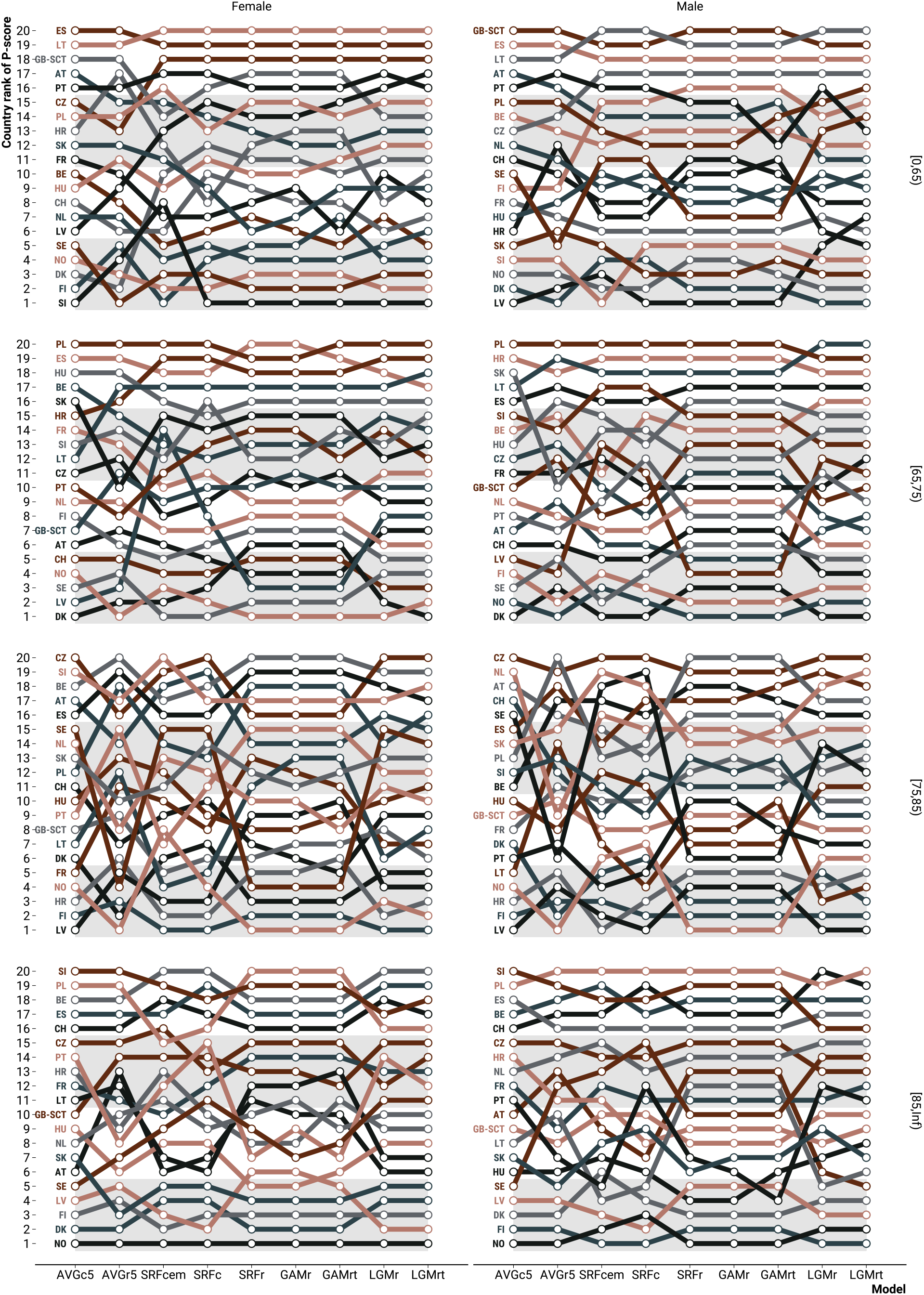
Within-stratum country ranking of excess death percentage during the year 2020 weeks 8 through 52 under 9 different models.

**Figure S.12:**
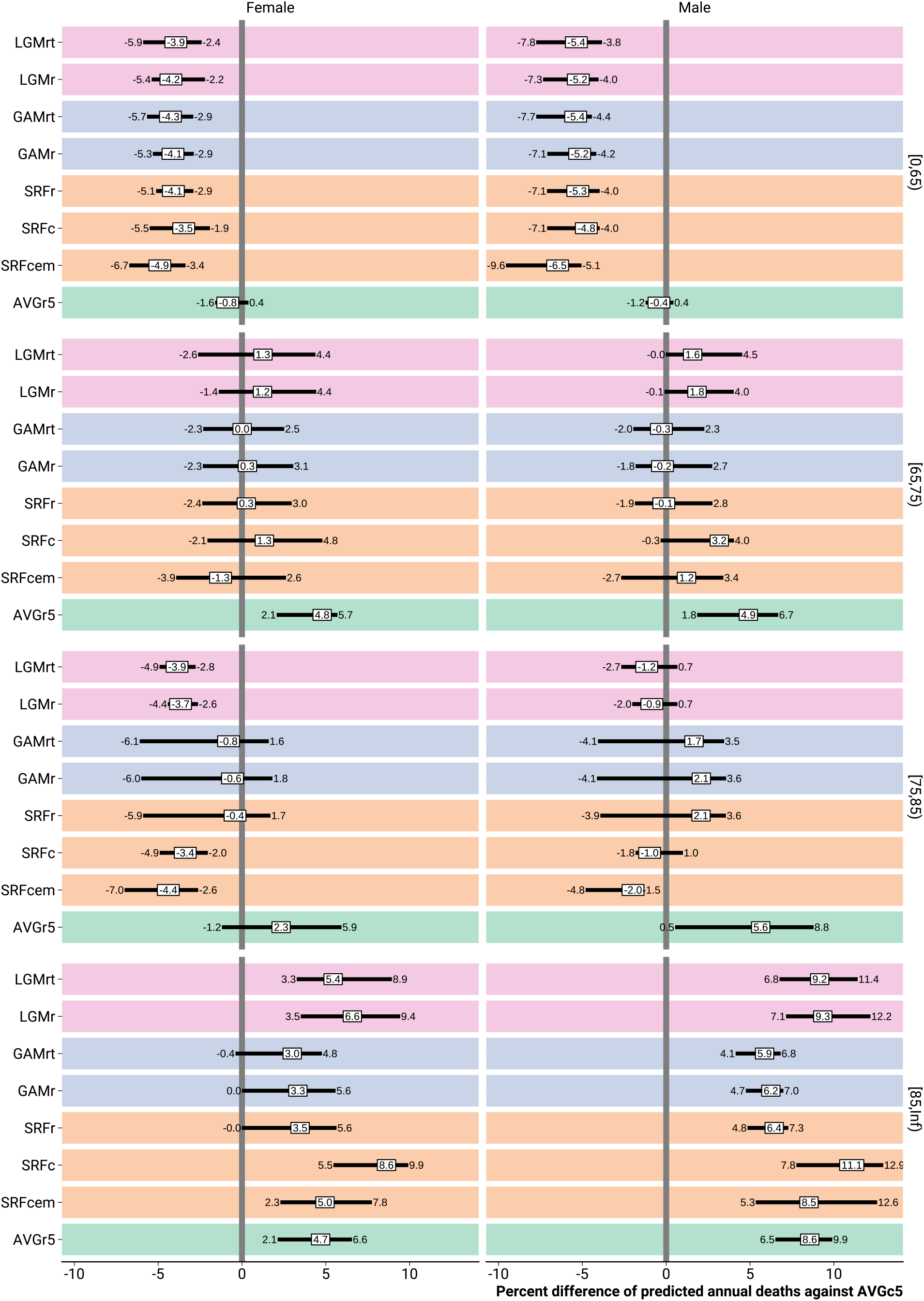
Within-stratum percent differences of predicted annual death counts from various models against the 5-year average weekly death count model prediction. *Note: The prediction differences were summarised across countries by the 0*.*25, 0*.*5, and 0*.*75 quantiles*.

**Figure S.13:**
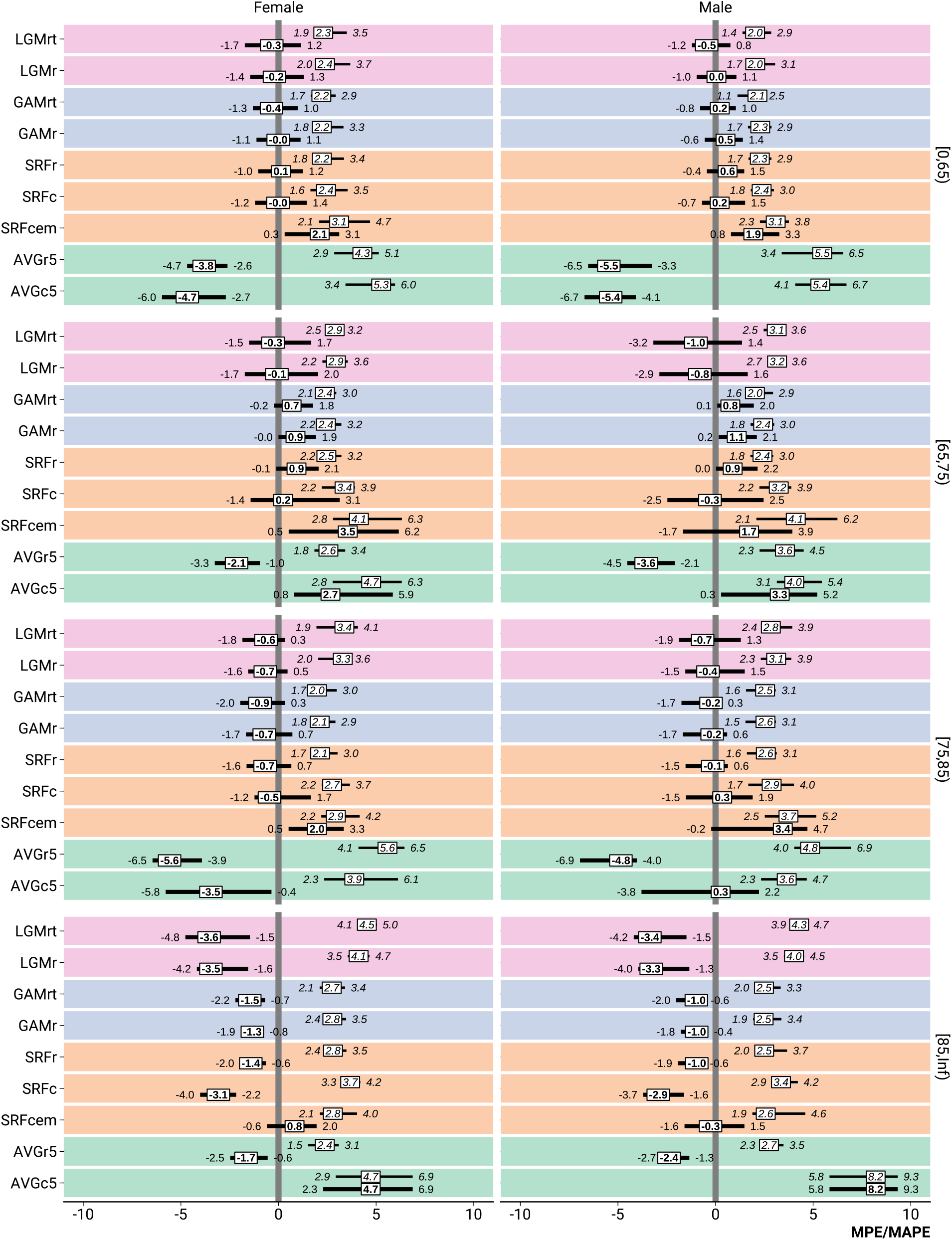
Within-stratum bias (MPE, bold) and error (MAPE, italic) by model when predicting annual death counts on test data. *Note: The error and bias measures were summarised across countries by the 0*.*25, 0*.*5, and 0*.*75 quantiles*.

